# Applying Multi-Theory Model (MTM) in Determining Intentions to Smoking Cessation among male Health Worker Smokers in Kabul, Afghanistan

**DOI:** 10.1101/2024.02.22.24303218

**Authors:** Mousa Bashir, Farkhondeh Amin Shokravi, Anooshiravan Kazem nezhad

**Author notes:** CORRESPONDING AUTHOR Farkhondeh Amine shokravi, No 227, Department of Health Education and Health Promotion, College of Health Sciences, Tarbiat Modares University, P.O. Box 14115-111 Nasr, Jalal Ale Ahmad, Tehran, Iran.

## Abstract

**Introduction:** Globally, smoking causes lung cancer and a wide range of acute and chronic diseases. A fourth-generation behavioral framework, the multi-theory model (MTM) of health behavior change was used to predict the initiation and maintenance of smoking cessation among health worker smokers.

**Methods:** By visiting different Health Centers, a convenience sample of male health worker smokers from west part of Kabul city, was invited to participate in this cross-sectional study. A valid and reliable 37-item MTM-based survey instrument was administered to the male participants who smoked. To explain smoking cessation behavior, stepwise multiple regressions were conducted. The entire value of the Cronbach alpha coefficient (α) of the subscales and the scale for the initiation of MTM variables was 0.80 and for the sustenance of MTM variables was 0.79.

**Results:** The study was completed by 170 participants. Participants were averaging 29.33 years of age (SD = 6.21). The average number of years smokers reported (SD = 4.7), was 5.6. Smoking cigarettes was the median (SD = 5.21), with 5.64 cigarettes consumed per day. Changes in the physical environment (+0.2225, P = 0.029) and behavioral confidence (+0.441, P = 0.014) were significant predictors of smoking cessation initiation. The intention to sustain smoking cessation behavior was significantly influenced by emotional transformation (β = 0.222, *P* = 0.017) and practicing for change (β = 0.217, *P* = 0.015).

**Conclusions:** There was moderate variance in smoking cessation behavior among health worker smokers in Kabul’s western part explained by two MTM constructs (behavioral confidence, physical environment) for initiation and two MTM constructs (emotional transformation, practicing for change) for maintenance. Smoking cessation behavior can be assessed using MTM both at the initiation and maintenance stages. It is important to develop future interventions using MTM constructs aiming to change smokers’ behavior in regard to quitting smoking.

## INTODUCTION

Smoking is one of the leading causes of preventable deaths worldwide [1]. The report of the World Health Organization estimated the prevalence of smoking in men will reach 30% by 2025 [2]. In the Middle East region and neighboring countries, the prevalence of smoking in men aged ≥ 15 years in 2020 is estimated as follows: Pakistan is 35, Iran is 20, and India is 17.8 [2]. World Health Organization reported the prevalence of smoking in Afghan men aged 15 years and older to be 35% in 2017. Also, this organization states that out of the five most common cancers among Afghan men, four cancers (stomach, esophagus, lung, and oral cavity) are attributed to smoking and other forms of tobacco use [3]. Smoking damages almost all the organs of the body, leads to countless diseases, and reduces the level of health in smokers [4]. According to Malaina et al.’s study, Coronary heart disease is linked to smoking, health outcomes will be improved by changing this behavior [5]. To change behavior, especially in the process of quitting smoking, health workers can play an essential role because they are both counselors and smoking cessation models for citizens [6]. Training programs are needed for health workers to increase their ability to actively support patients with smoking cessation techniques [7, 8]. Two important results from the studies show that if health workers are not smokers themselves, they are more successful in persuading smokers to quit smoking, as well as smokers who are supported and counseled by healthcare providers, than those who do it themselves have more chances to quit smoking [9]. In addition, studies show a relatively high incidence of smoking among health workers, especially in some countries such as Iraq (26.5 percent) [10]. Also, research has shown that health workers have received little formal training to quit smoking. (7.3 percent) [11]. Also, they had little knowledge about smoking cessation strategies [12]. Meaning that these staffs are not good role models for their patients [13]. It has been shown that public health interventions to promote smoking cessation are effective, but the potential for applying public health intervention across community is limited. There is inadequate predictive power in previous health behavior models used to describe smoking cessation, and long-term behavioral change cannot be evaluated using those models [14]. As a health behavior theory, the multi-theory model employs a fourth-generation framework for simultaneous prediction and modifying health behaviors over a long period of time [15, 16].

The MTM is an efficient and effective theory for assessing initiating as well as sustaining the prevention, reduction, and cessation of smoking [14], [17-20]. Moreover, Health behaviors in a wide variety of populations have been explained using the MTM [21-32]. The initiation of behavior model has three constructs: 1. Participatory dialogue (advantages offsetting the disadvantages of the health behavior change), 2. Behavioral confidence (beliefs that one can perform the behavior change), 3. Changes in the physical environment (having resources at one’s disposal for the behavior change) [15, 33] (Figure 1). Constructs can be divided into three types to maintain behavior: 1. emotional transformation (translating feelings into goals for the behavior change), 2. Practice for change (creating a habit of transformation and making it a way of life), 3. Changes in the social environment (obtaining social support to help one maintain the health behavior change) [15, 33] (Figure 2). In Afghanistan there have not been many studies about tobacco, an application of the Multi-Theory Model was the purpose of this study in determining intentions toward smoking cessation among Afghan male health worker smokers.

**Figure 1:**
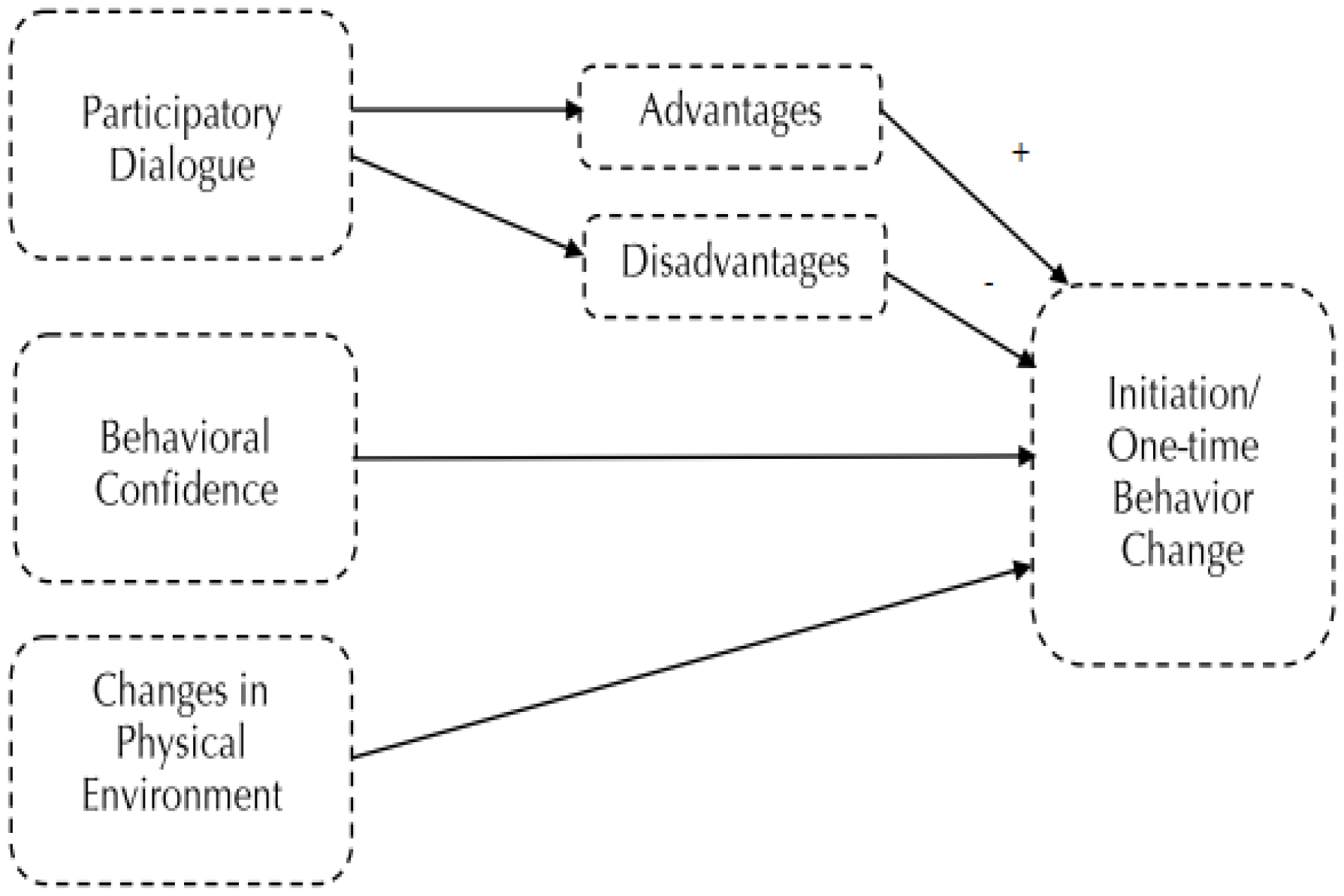
Presents constructs in the initiation of smoking cessation in the multi-theory model

**Figure 2:**
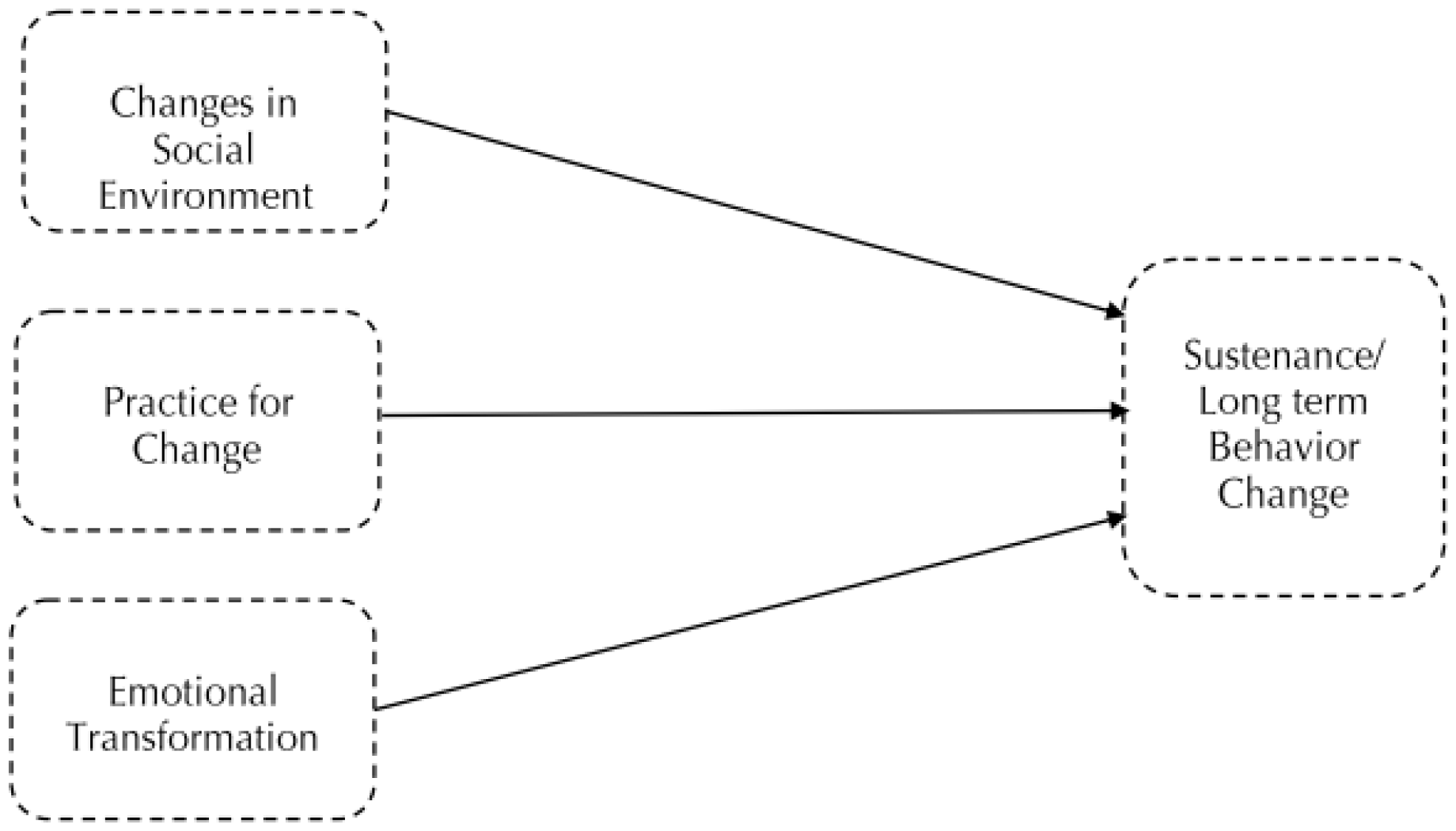
Presents constructs in the sustenance of smoking cessation in the multi-theory model

## METHOD

A cross-sectional research design was used for this study. The research was conducted in the west of Kabul, Afghanistan, participants in the study were male health worker smokers. Data was collected from April to July 2021. Participants were included if they smoked one or more cigarettes during the past seven days, 20 years of age or older. In this study, convenience sampling was applied. For stepwise multiple regressions, a minimum sample size of 170 participants is required if the alpha is 0.05, the power is 0.90, effect size equal to 0.1, response rate 80, and each model includes three predictors and five covariates suing Gpower 3.1 software. [34].

In this study, the variables that were independent were the components of the initiation model (participatory dialogue, behavioral confidence, changes in the physical environment) and the components of the sustenance model (emotional transformation, practice for change, and changes in the social environment). In both models, intentions to initiate and sustain smoking cessation behavior changes were dependent variables.

### Instruments

In this study a self-report MTM based questionnaire consisting of 37 items was used as an instrument. A person’s smoking status is assessed in the first eight questions of the questionnaire (number of cigarettes smoked per day, years of smoking, current smoking status), as well as demographic characteristics (their educational level, age, income, employment status). Furthermore, the MTM model constructs were assessed using 29 items associated with the two models the initiation model including participatory dialogue, behavioral confidence, changes in the physical environment and sustenance models including emotional transformation, practice for change, and changes in the social environment [14]. The original questionnaire was translated and then re-translated.

### Smoking cessation: Starting and Maintaining

It is important to understand that initiation of smoking cessation includes deciding to quit smoking, while sustenance relates to the achievement of abstinence [35]. In both items, the following ratings were given: not at all likely (0), somewhat likely (1), moderately likely (2), very likely (3), and completely likely (4) [35].

The initiation model is comprised of three constructs that are measured by 19 items. Participatory dialogue, the first item, Provides information on how to quit smoking and its advantages and disadvantages. Smoking cessation benefits are discussed in five questions in advantages (i.e. being healthy, able to save money, getting sick less often, smelling better, & enjoy life more) and disadvantages also have five questions related to difficulties of smoking cessation (i.e., not able to relax as well, not able to socialize as well, miss it, not be able to overcome the urge and loss friends). Following is a list of advantages and disadvantages: Never (1), almost never (2), sometimes (3), fairly often (3), and very often (4). A possible total score ranged from 0 to 20. According to this hypothesis, higher scores for advantages and lower scores for disadvantages were related to the initiation of smoking cessation or behavior change. By subtracting disadvantages from advantages, the participatory dialogue score was obtained. Its value ranged from -20 to +20 [11, 12]. The second item is behavioral confidence, a five-question assessment is used to determine whether a person is confident about quitting smoking. i.e. ability to quit smoking this week, ability to quit smoking this week and complete all work-related tasks, ability to quit smoking this week and feel relaxed, ability to quit this week without getting anxious, and ability to quit smoking this week without getting withdrawn symptoms. In each item, the responses were rated as follows: Not at all sure (0), slightly sure (1), moderately sure (2), very sure (3), and completely sure (4). 0 to 20 was the range of possible total scores. Those with a higher score are more likely to initiate smoking cessation [35]. There are three questions regarding the physical environment that contributes to quitting smoking in the third item, which refers to changes in the physical environment, i.e. ability to get rid of all cigarettes from one’s environment this week, the ability not to buy cigarettes this week, and ability to substitute smoking time with something else this week. The response to each item was rated as follows: Not at all sure (0) to completely sure (4). The possible total score range was 0 to 12. High-scoring individuals are more likely to initiate a smoking cessation program [35]. There were ten items used to assess the constructs of the sustenance model. The first item is emotional transformation, which consists of three questions related to emotions that providing smoking cessation assistance, i.e. the ability to direct one’s emotions/feelings to the goal of being smoke-free every week, the ability to motivate oneself to be smoke-free every week, and ability to overcome self-doubt in accomplishing the goal of being smoke-free. The following grading system was used for each item: From not at all sure (0) to completely sure (4). In total, 12 possible scores were possible. A higher score indicates a greater likelihood of sustained quitting [35]. The second item is practice for changes, which is broken down into three questions that you can quit smoking with these tips: i.e. Ability to keep a weekly self-diary to monitor smoking urges, when faced with barriers, be smoke-free every week and change the plan to be smoke-free every week if problems arise. Responses to each item were rated as follows: From not at all sure (0) to completely sure (4). The range of possible scores is 0 to 12. Those with higher scores are more likely to sustain their smoking cessation efforts [35]. As the third item, changes in the social environment, to determine whether family members, friends, and health care providers are likely to support you, three questions must be asked i.e. Ability to get weekly support from a family member to quit smoking, Ability to get assistance from a friend each week, try not to smoke and ability to receive assistance from a health professional to quit smoking every week. Responses to each item were rated as follows: From not at all sure (0) to completely sure (4). Between 0 and 12 is the possible total score. Higher scores indicate a greater chance of success in quitting smoking [35].

### Ethical aspects

The Research Ethics Committee of Tarbiat Modares University granted ethics approval for this study (IR.MODARES.REC.1399.256). Participants were initially informed of the objectives and all phases of the process were conducted in strict confidentiality. We collected no personal information from participants and the participation was completely voluntary. A written informed consent was obtained and the correct information was requested.

### Validity and reliability

In the present study, a questionnaire created in 2017 by Professor Sharma and colleagues entitled “ Quitting smoking by applying a new theory: Change in behavior using multi-theory models “ was used. Its validity and reliability have been measured and established [35].

### Statistical analysis

A descriptive analysis was performed on all study variables (Tables 1 and 2). As appropriate, we calculated Spearman’s r to evaluate the variables related to demographics (covariates) and dependents variables (Smoking cessation behavior initiation and maintenance). Since there were two dependent variables, in order to determine how predictive MTM constructs are, we conducted two stepwise multiple regression models in two blocks apart from factors influencing demographics (Tables 3 and 4). Each model began with block 1 entering demographic variables in bivariate analysis. Every MTM construct was added to block 2 for each model. A priori, 0.05 was set as the statistical significance level. The IBM SPSS V.25 is sued for analyzing the data.

**Table 1.**
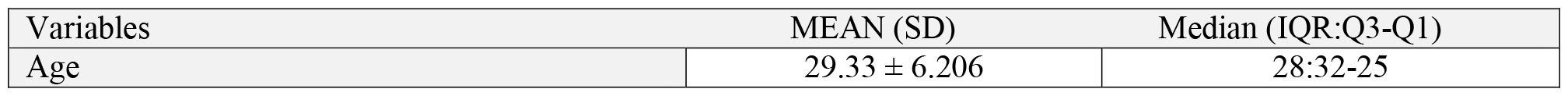
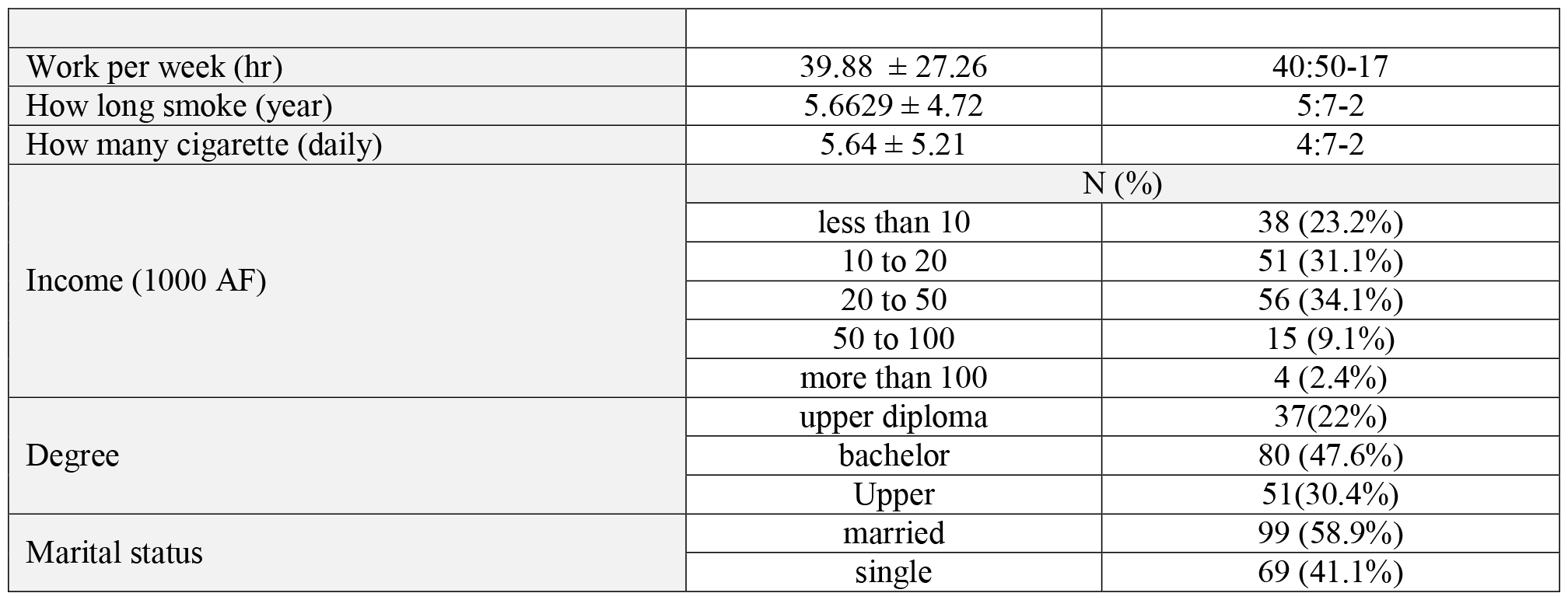
Socio-demographic characteristics of the participants.

**Table 2.**
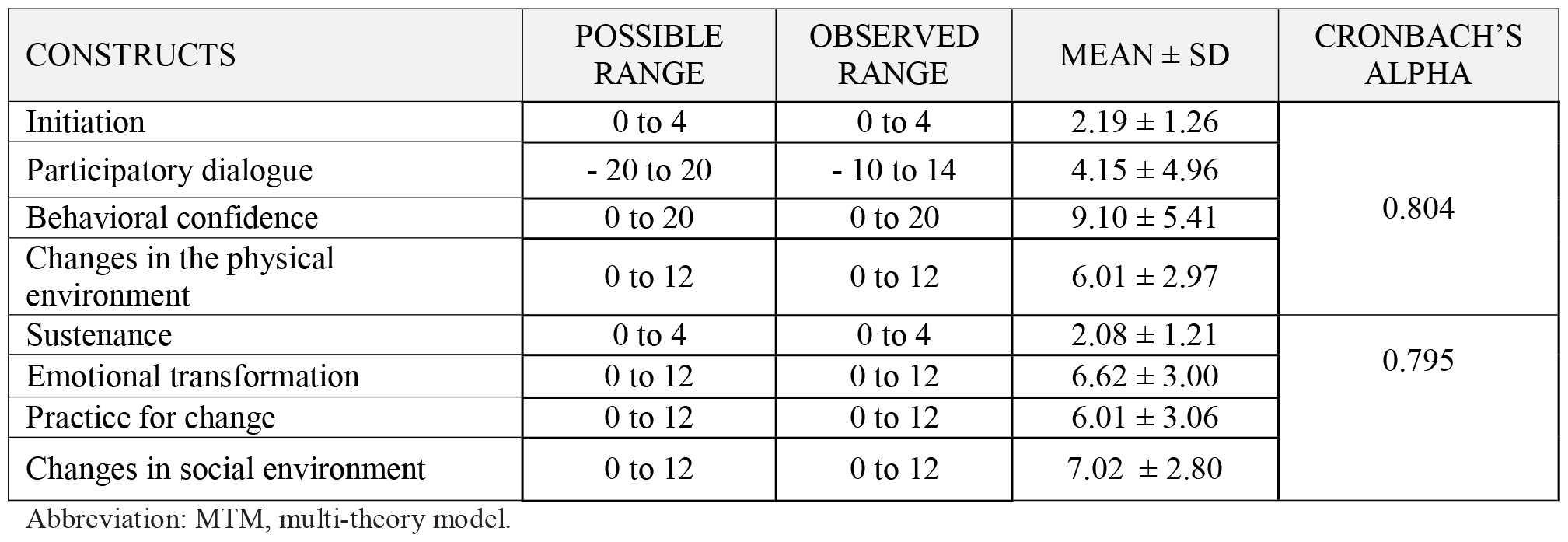
Descriptive statistics of constructs of MTM (n = 170).

**Table 3.**
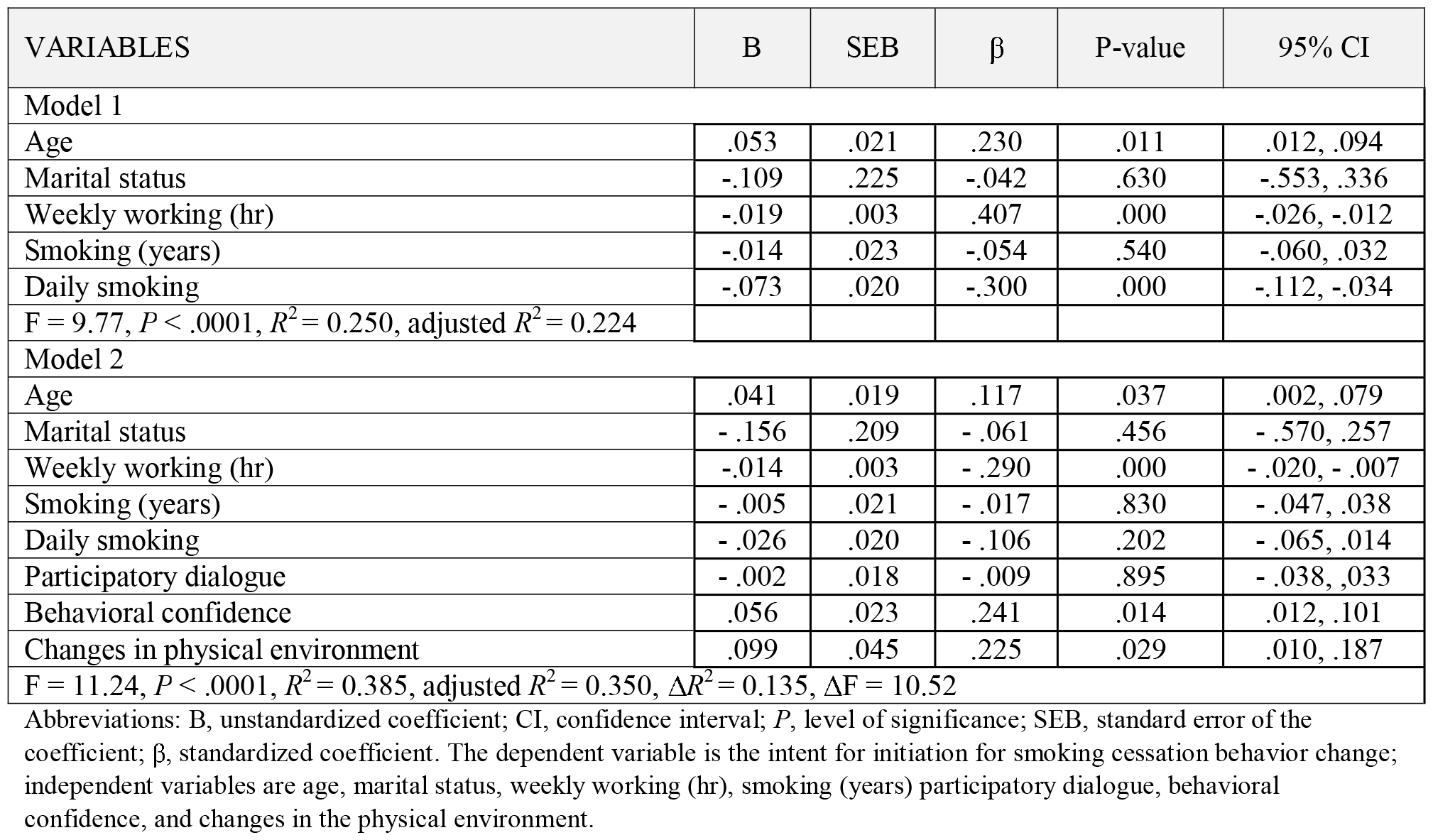
Stepwise multiple regressions predicting initiation for smoking cessation (n = 170)

**Table 4.**
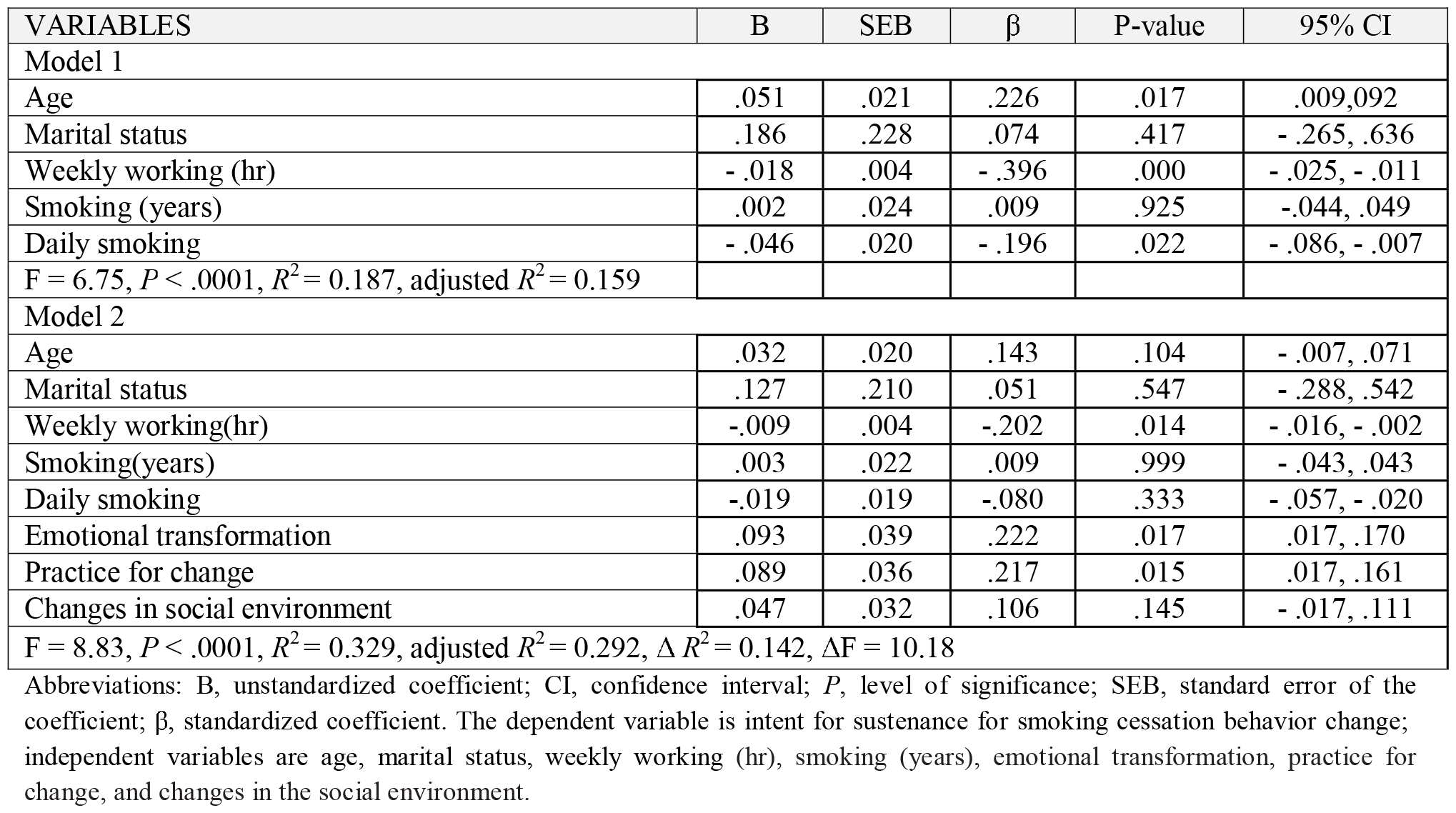
Stepwise multiple regressions predicting sustenance for smoking cessation (n = 170)

## RESULTS

Participants meeting the inclusion criteria (aged 20 and over and smoked one or more cigarettes during the past seven days) totaled 170 and filled out the questionnaire on paper. Among the participants, the mean age was 29.33 years (SD = 6.21). According to the participants, they smoked for an average of 5.6 years (SD = 4.7), Cigarettes smoked per day averaged 5.64 (SD =5.21). Participants typically had an income between 20000 and 50000 AF and most had a bachelor’s degree. Table 1 provides comprehensive demographic information. All MTM constructs are described in Table 2 as well as their descriptive statistics and reliability calculations. Based on a 5-point scale, there was a mean score of 2.19 (SD =1.26) for the intention of initiation. In the initiation model the participatory dialogue had a mean of 4.15 units (SD = 4.96, range = –20 to 20), the behavioral confidence had a mean of 9.10 units (SD = 5.41, range = 0-20), the changes in physical environment had a mean of 6.01(SD = 2.97, range = 0 to 12). Based on a 5-point scale, there was a mean score of 2.08 (SD =1.21) for the intention of sustenance. In the sustenance model, the emotional transformation had a mean of 6.01 (SD =3.06, range = 0-12), the practice for change had a mean of 7.02 (SD =2.80, range = 0-12), the changes in social environment had a mean of 6.62 (SD=3.00, range = 0-12).

The first part of the table (model 1) demographic factors were found to be predictive of smoking cessation initiation, age (β = 0.230, P = .011), weekly working hours (β = 0.407, P = .000), and daily smoking (β = –0.300, P = .000) were significant, F = 9.77, P = .0001, R^2^ = 0.250. The second part of the table (model 2) an increase of 0.135 in R^2^ was observed after adding the initiation model MTM constructs. A statistically significant result was obtained for the overall model, F = 11.24, P < .0001, R^2^ = 0.385, adjusted R^2^ = 0.350. In model 2, smoking cessation initiation behavior was significantly predicted by behavioral confidence (β = 0.241, P = .014) and changes in physical environmental (β = 0.225, P = .029). It states, behavioral confidence (β = 0.241, P = .014) and changes in the physical environment constructs (β = 0.225, P = .029) were significant predictors of initiation for smoking cessation behavior in model 2. A stepwise multiple regression analysis of the sustenance model is presented in Table 4. The first part of the table (model 1) demographic factors were found to be predictive of smoking cessation sustenance, weekly working (hr) (β = - 0.396, P = .000), daily smoking (β = - 0.196, P = .022), P = .0001, R^2^ = 0.187, adjustment R^2^ stands at 0.159. A statistically significant increase in the R^2^ value of 0.142 was observed with the addition of the sustenance model MTM constructs to model 2. A statistically significant result was obtained for the overall model, R^2^ = .267, the adjusted R^2^ is 0.292, and P < .0001. A significant predictor of the sustenance of cessation behavior with regard to smoking was the level of emotional transformation (β = 0.222, P = .017) and practice for change (β = 0.217, P = .015).

## DISCUSSION

This study was conducted to examine if the MTM constructs can predict both the intention to initiate smoking behavior and the intention to maintain it among male health workers in the western part of Kabul.

Many studies have been conducted on smoking behavior based on the MTM theory including smoking cessation, tobacco cessation, water pipe smoking, cigarette smoking, substance use behavior, [14, 17, 18,19,20,35, 36, 37]. The initiation and sustenance of behavior change are substantiated in predicting health behavior changes by the finding of these studies. An average of good intentions was reported for initiation (i.e., quitting in the near future) and sustainability of smoking cessation (i.e. sustaining smoking cessation for the foreseeable future) among the participants.

There was an increase in the R^2^ of both the initiation regression model and the sustenance regression model after the addition of MTM constructs without considering the demographic covariates. The behavioral confidence of smokers as well as changes in their physical environment were significant predictors of initiation of smoking cessation behaviors. Sustenance for smoking cessation behavior was significantly predicted by emotional transformation and practice for change. It has been determined that 35.0% of the variance in smoking cessation initiation is attributed to age, weekly working hours, daily smoking, behavioral confidence, and physical environment changes, Nahar et al. 2019, showed 23.6% of the variance is attributed to daily smoking, participatory dialogue, behavioral confidence [14], Sumitra SHARMA et al. 2020, reported 48% of the variance in the initiation of smoking cessation is explained by behavioral confidence and changes in the physical environment [17]. Studies conducted in two countries, Nepal and Afghanistan, showed that behavioral confidence and changing the physical environment are more effective in predicting the initiation of behavior change and it is marked with a high percentage. However, the study conducted in the United States found that only behavioral confidence was effective in initiating behavior change. It is likely that the difference in the percentage of Predicting smoking cessation initiation behavior in the United States (23.6%) versus Nepal (48%) and Afghanistan (35.0%), could be related to differences between study contexts. Additionally, a positive correlation is found between both MTM variables and the initiation of smoking cessation.

According to our study, participatory dialogue did not predict smoking cessation initiation. The studies of Bashirian et al. 2019, and Nahar et al. 2019, however, have concluded that participatory dialogue is a significant predictor of water pipe smoking and smoking cessation among smokers [14, 19]. Additionally, previous studies have demonstrated that initiating behavior change in other health behaviors is significantly predicted by participatory dialogue, including smoking cessation, water pipe smoking, portion size, physical activity, and mammography screening [14, 19, 22, 24, 32]. Studies related to MTM have demonstrated that participatory dialogue is linked to other health behaviors including behavioral confidence is operationalized in the MTM as a person’s confidence to engage in a behavior change in the future and was derived from Bandura’s self-efficacy and Ajzen’s perceived behavioral control [35].

There has been a significant relationship between perceived behavioral control and self-efficacy and smoking cessation in previous research. Studies show that smoking cessation rates are higher with higher self-efficacy [8]. Research suggests that perceived behavioral control and self-efficacy exert in the effort to diminish smoking behavior, behavioral modification needs to be emphasized as a tool to enhance the level of success in quitting smoking [8, 14, 38]. Previous studies on smoking cessation and water pipe smoking showed that behavioral confidence was an important predictor of smoking cessation and water pipe smoking reduction, the present study also confirms these findings [14, 17, 19]. Previous studies on the constructs of MTM and other behavior-related health measures, for example Physical activities, consumption of sugar-sweetened beverages, binge drinking, eating habits, sleep, outdoor nature contact behavior, and mammography screening, behavioral confidence has consistently been an important predictor of behavior change initiation [22, 23, 25, 27-29, 31, 32, 39].

Physical environment changes significantly contributed to smoking cessation, according to our study. It means that we are able to remove all cigarettes from the environment, to be able to refrain from purchasing cigarettes, this is a concept derived from Bandura’s social cognitive theory which is referred to as substitutability of activities in substituting smoking time with something else [14, 19, 21, 22, 24, 27, 31, 31-38, 40-43]. Changing the physical environment was found to be an important predictor of a person’s decision to quit smoking in the present study; which is consistent with the study of smoking cessation in Kathmandu Metropolitan City, Nepal [17]. Initiation of other health behaviors is also predicted by MTM, including overdrinking, beverages sweetened with sugar, physical activity, outdoor nature contact behavior, SAVOR (Sisters Adding Fruits and Vegetables for Optimal Results), HPV vaccination, sugar-sweetened beverages, and mammography screenings[17, 23, 25, 28-32, 38]. Different health behaviors across countries are often influenced by environmental factors in combination with individual, family, and community factors [44].

Only practice for change and emotional transformation were significant predictors in the sustenance model, these factors explain 29.2% of the variance, Nahar et al. 2019, reported only emotional transformation was a significant predictor and explained 23.3% of the variance [14] and Sumitra SHARMA et al. 2020, showed 54% of the variance is explained by emotional transformation [17]. In these three studies, emotional transformation played a key role in maintaining behavior change, again in Nepal and Afghanistan, this role is more prominent than in the United States. It is revealed that maintaining behavior change is as effective as initiating behavior. Sustenance is positively correlated with both variables. Emotional intelligence is essential to the transformation of an individual’s emotions and consequently can be used in the MTM to direct emotion in a way that will help the smoker quit and change their behavior by overcoming uncertainty [35]. There is a positive correlation between emotional transformation and the sustainability of studies such as smoking cessation [14, 17], tobacco cessation [18], water pipe smoking [19], Smoking cessation in youth [20], and vaping quitting behavior [45].

Smoking cessation sustainment was significantly predicted by practice for change in our study. The practice for change has been identified as a significant predictor of behavior sustenance in previous studies exploring MTM constructs such as water pipe smoking and cigarette smoking in youth [19, 20]. MTM has also been found to be associated with the practice for change and sustainability of other health behaviors in previous studies utilizing the MTM, such as Outdoor Nature Contact Behavior, SAVOR (Sisters Adding Fruits and Vegetables for Optimal Results), physical activity, vaccination, sugar-sweetened beverages, mammography screening [25, 28-32].

Smoking cessation sustenance was not significantly predicted by changes in the social environment in this study. An investigation by Abasi et al., titled “Cigarette smoking in Youth, using the MTM”, has revealed that social environment plays a significant role in the sustenance of cigarette smoking among youth [20]. Using the MTM to examine the social environment, other health behaviors were found to be significantly influenced by the social environment, including binge drinking, sleep, physical activity, dietary behavior, outdoor nature contact behavior, SAVOR, mammography screening [22, 23, 27-29, 32, 38]. Although this study revealed a negative effect, other studies have indicated positive effects on smoking cessation behavior in the form of family and peer support [32, 46].

### Strengths and Limitations

The application of new theory of health behavior change (MTM) is strength of this study. However, this study has limitations that the authors would like to acknowledge. In this study, due to target population study limitation, convenience sampling method was used for male health workers who smoke. Therefore, the findings cannot be generalized beyond the sample population of the study. Also there is a need for future research to use randomized controlled designs to conduct interventional studies. The current study was restricted to only male health worker smokers. Future studies can be planned to investigate both genders. Finally, in this study our sample was from the west region of the Kabul city, so it is not representative of all regions, which may have influenced the results.

## CONCLUSIONS

Health workers in the western part of Kabul showed significant predictor relationships with two MTM constructs for initiating change and two MTM constructs for maintaining change. The MTM was shown to be accurate at both predicting the initiation of smoking cessation and its sustainability according to the findings of this study. In this study, MTM constructs were successful in initiating and maintaining behavior change through the development of smoking cessation interventions. Based on the MTM, smoking cessation interventions should incorporate behavioral confidence, build upon an individual’s change in the physical environment to initiate smoking cessation, and encourage individuals to sustain their smoking cessation efforts by enhancing their emotional responses to behavior change. For the development of tobacco cessation interventions in the future more studies are needed to determine whether the MTM constructs are predictive in other, more diverse and randomized samples to improve generalizability.

## Data Availability

All data produced in the present study are available upon reasonable request to the authors

## Acknowledgments

The author would like to thank Professor Manoj Sharma a theorist of Multi Theory Model of health behavior change (Department of Social and Behavioral Health, the University of Nevada, USA) for his guidance in providing the MTM-based proposal, during the thesis implementation and as a final reviewer of this article.

## Declarations

### Ethics approval and consent to participate

The study was conducted according to the guidelines of the Declaration of Helsinki and approved by the Ethics Committee of IR.MODARES.REC. 1399.256 ethical code in Tarbiat Modares University Ethical Committee. And informed consent was obtained from all subjects involved in the study.

### Consent for publication

Consent for publication not applicable.

### Availability of data and materials

Data are confidential and are not available upon request.

### Authors’ contributions

MB. And FSH. Wrote the main manuscript text and AK. Prepared figures. All authors reviewed the manuscript.

### Author details

1. Department of Health Education and Health Promotion, School of Medical Sciences Tarbiat Modares University Tehran.Iran
2. Department of Bio statistical, School of Health Sciences, Tarbiat Modares University, Tehran, Iran

